# TIPS (The Trans-Tasman Internet-delivered Prevention of (youth) Suicide) Study: Protocol for a Randomised Controlled Trial of Mobile Health Interventions (four apps) to help young people reduce suicidal ideation

**DOI:** 10.1101/2025.11.27.25341169

**Authors:** Nicola Ludin, Michelle Torok, Lauren McGillivray, Quincy Wong, Sarah Fortune, Inge Meinhardt, Hiran Thabrew, Tania Cargo, Sarah Hetrick

## Abstract

**Introduction:** Suicidal ideation is common in young people and increases the risk of suicide. Effective interventions that are relevant and accessible to young people, so-called digital natives, are urgently required. There are key questions regarding the cross-cultural efficacy of suicide prevention apps for scalability.

**Methods and analysis:** This online four-arm parallel randomised controlled superiority trial will enrol 1480 young people aged 16-24 with current suicidal ideation in New Zealand and Australia. Participants will be randomised to one of three therapeutic apps developed in different countries, Tune In, Bro and LifeBuoy, or to My Mood (attention control). The primary outcome is suicidal ideation severity at 30-day and 90-day post-baseline; secondary outcomes include mental well-being, engagement and acceptability. Emotional regulation will be examined as a mediator of change in suicidal ideation. All outcomes are measured by self-reported scales incorporated in an online questionnaire. Acceptability of the apps for rangatahi Māori (Indigenous youth, New Zealand) will be explored via semi-structured interviews. Linear mixed models with repeated measures analyses, employing maximum likelihood estimation, an appropriate covariance structure, and consideration of site effects, will be undertaken. Examination of individual app intervention effects in New Zealand and Australia will highlight the effects of apps developed for a different country.

**Ethics and dissemination:** Approval was obtained (26 February 2025) from the Health and Disability Ethics Committees (Ministry of Health ref 2025 EXP 21500). It was registered in the Australian New Zealand Clinical Trials Registry ACTRN12625000349448 23/04/2025; recruitment started on July 14, 2025 (594 recruited to date), and is expected to be complete by June 2026. Participants provide informed consent online. Trial results will be submitted for publication in peer-reviewed journals, shared on relevant websites, and via presentation at international scientific conferences; IPD will only be shared if requested and subsequent to review.

**Article Summary:**

- This is one of the largest online trial of suicide prevention apps in young people ever undertaken.
- This trial will test several different suicide prevention apps in a four-arm parallel superiority randomised trial design.
- The trial will specifically examine the cross-cultural efficacy of apps by testing the effectiveness of an app developed in a country different from where it is being tested.

**Strengths and limitations:**

- There is a risk of fraudulent behaviour in online trials; in this trial, this primarily consists of providing false information to ensure eligibility. Targeted social media advertising (for age and location) for recruitment will minimise risks. SMS verification ensures protection against bots and multiple accounts.

## Introduction

### Background and rationale

Suicidal ideation (thoughts about, or plans of, ending one’s life) doubles the risk of death by suicide (1). Internationally, lifetime rates are as high as 24% in those aged up to 25 years (2, 3). In Aotearoa, New Zealand, the 12-month prevalence rates in adolescents have increased from 15.3% in 2012 to 20.8% in 2019 (4). This equates to around one in five young people experiencing suicidal ideation each year. Rangatahi Māori (Māori youth, the indigenous people of Aotearoa) are twice as likely to be affected (5), similar to other countries with Indigenous populations, like Australia, where Aboriginal and Torres Strait Islander youth are also more impacted (6).

Addressing suicidal ideation is a critical strategy to address youth suicide rates (4). As with Indigenous populations globally, in Aotearoa, New Zealand rangatahi Māori face disproportionately higher rates of suicide compared to their non-Māori peers (4, 7–9) due to ongoing intergenerational impacts of colonisation, institutional racism, and cultural trauma (10). The most recent rate per 100,000 of suspected suicide was 26.3 for Māori aged 15 to 24, compared with 15.6 per 100,000 for non-Māori (11). This represents a breach of Te Tiriti o Waitangi (Treaty of Waitangi) obligations by the Crown (12) and must be redressed.

Face-to-face services are valuable, but they are not suitable for everyone, and are often experienced as culturally unsafe or inappropriate, particularly for rangatahi Māori. Many youth who experience suicidal ideation do not have a severe mental illness, so are unlikely to meet the threshold for entry to public mental health services (13). There are many other well documented reasons they do not access adequate or effective public mental health or other support services (14–16), particularly specific populations like Indigenous (17, 18), rainbow (19, 20) and for those in low- to middle-income countries or living rurally or in more isolated areas (21–23).

Today, most young people look for support and mental health information online, including via social media and mental health applications (apps) (24–27). Youth born between the late 1990s and late 2010s, so-called digital natives, have not known anything but the everyday use of digital technologies and social media, which are integrated into their daily lives with little perceived difference between their online and offline worlds (28–30). There is an opportunity to provide evidence-based interventions within this context, including integration into digital platforms as highlighted by a recent study in Australia (31) of young people, policymakers, and social media platform professionals. A systematic review in 2020 highlighted promising results for such tools; however, few had been rigorously tested in randomised controlled trials (32). Since then, trials have showed positive results for young people hospitalised for suicidal ideation in the US (33, 34), and for community-based young people in Australia with suicidal ideation (35, 36).

A key question is whether digital interventions for suicidal ideation are effective regardless of their origin, or whether different jurisdictions require locally developed interventions. In areas like youth depression, research has shown that SPARX, a digital CBT intervention developed and effective in Aotearoa New Zealand (37), was also effective when trialled in Australia (38).

There is a clear need to examine the cross-cultural applicability of digital interventions for young people with suicidal thinking. For example, while LifeBuoy has been shown to be effective in large RCTs undertaken in Australia (35, 36), it is unclear if this app will resonate and be effective with Aotearoa, New Zealand youth. Similarly, two apps developed in Aotearoa New Zealand, Bro and Tune In, may or may not be effective for Australian youth. Understanding this is critical for decisions regarding scalability and promotion, and particularly for ensuring equity of outcomes for populations disproportionately impacted by suicide.

### Objectives

#### Primary aim: Efficacy of therapeutic apps compared with an attention control

To evaluate whether each of three therapeutic apps, Tune In, Bro and LifeBuoy, lead to greater reductions in suicidal ideation severity at 30-day and 90-day post-baseline in young people aged 16 to 24 years with current suicidal ideation, relative to My Mood (attention control mood monitoring app) in a randomised control trial in Aotearoa, New Zealand and Australia.

#### Secondary aims

1. To determine whether each intervention app will result in greater increases in mental wellbeing than control at 30-day and 90-day post baseline;
2. To determine if the intervention apps will be more engaging and acceptable to users than the control app;
3. To determine if improved emotional regulation mediates change in suicidal ideation at 30-day and 90-day post baseline;
4. To explore, qualitatively, the acceptability of the apps for rangatahi Māori.

## Methods

### Trial Design

This is a four-arm parallel randomised controlled superiority trial where three intervention apps will each be compared to one matched attention control app, rather than to each other. See Figure 1 for study flow. This protocol adheres to the SPIRIT guidelines (39).

**Figure 1.**
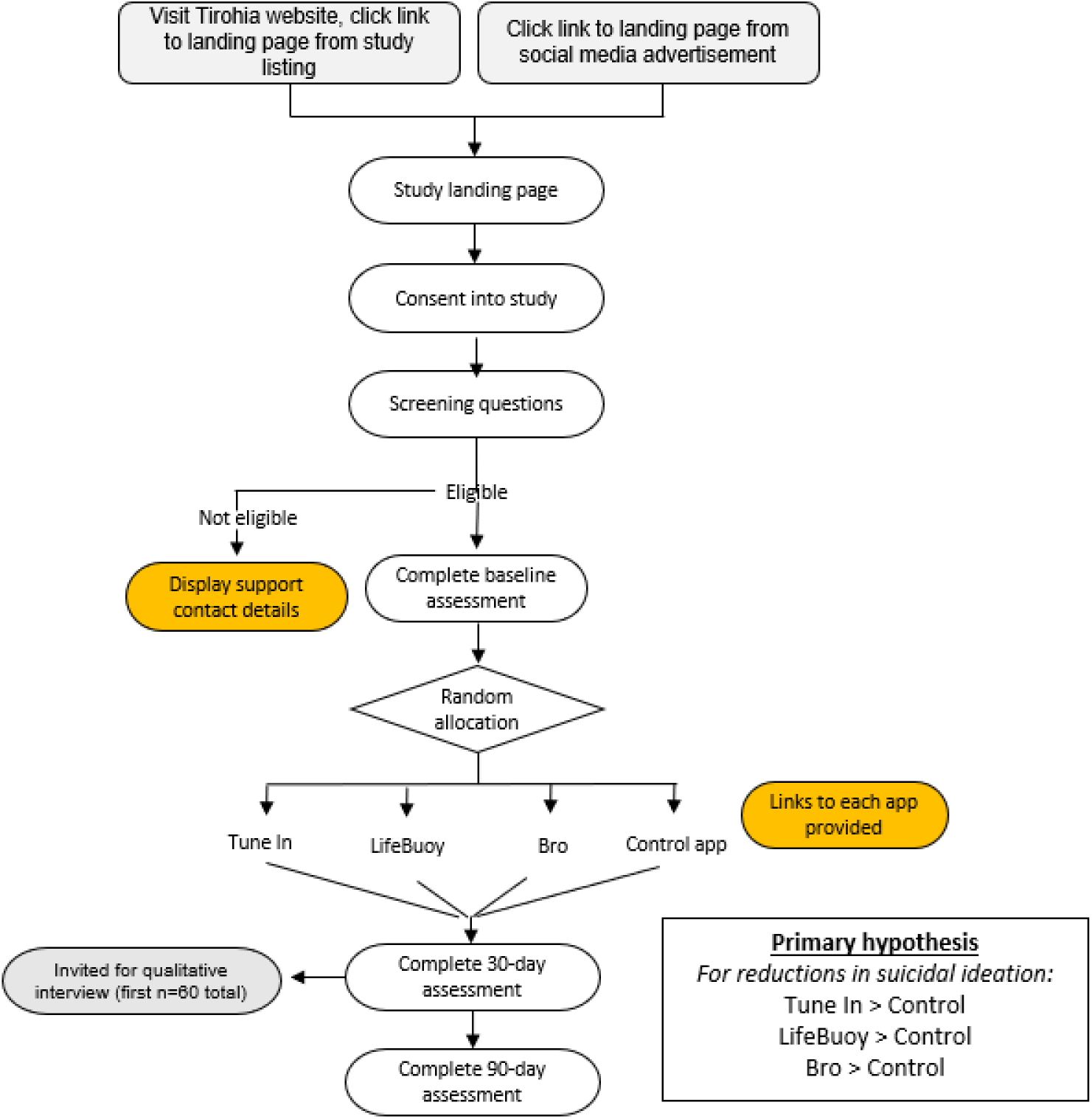
TIPS trial flow diagram

### Participants

Participants will be between 16 and 24 years of age and (1) live in either Australia or Aotearoa, New Zealand; (2) own or have access to a smartphone, minimum version iOS-V.13 or Android V.7 (3) have current (last month) suicidal ideation measured by the Suicidal Ideation Attributes Scale (SIDAS) [36], with or without self-harm. Participants will not be eligible if they have used the app they are allocated in a previous trial. There are no other exclusion criteria.

This online trial will be run on Tirohia, (40), a secure, configurable Web portal and suite of Web services that facilitates concurrent field trials. The primary recruitment method is via social media advertising. In Aotearoa New Zealand, recruitment will also include printed posters displaying the QR code/URL placed in community youth venues, schools and universities. Social media advertising and posters will be co-designed by social media-experienced youth researchers and a Rōpu Rangatahi Kairangahau Tipu Māori (Māori youth advisory group) to ensure the inclusion of rangatahi Māori. Social media content will also be shared via trusted partner organisations (e.g. Youthline, Mental Health Foundation, Voices of Hope).

Participants self-select into the trial based on the eligibility criteria included in the co-designed advertising and are directed to the TIPS study landing page, where they can view the Participant Information Sheet (PIS), and are advised to contact the researchers if they have questions. The PIS can be downloaded at the time of reading and from the study landing page. Informed consent will be captured digitally on the landing page via an electronic consent form (mandatory checkbox and timestamped submission). After consenting and completing the baseline assessment, participants are assigned a unique globally unique identifier (GUID) and presented with the app download.

### Interventions

Young people will have uninterrupted access to their app until the final participant has completed the 90-day follow-up. The apps are Tune In, Bro, LifeBuoy, and the control, My Mood. They include a curated collection of support resources and helpful links relevant to common mental health concerns in both Aotearoa, New Zealand and Australia.

### Tune In

Tune In was co-designed with 17 young people (12 females and five males; five NZ European, seven Māori, and five Pasifika). It is a goal-setting app that facilitates behaviour change in a culturally responsive way. Goal setting enhances emotion regulation by providing a sense of control and self-efficacy and promoting positive coping via planned engagement in strength-based behavioural strategies to achieve a personally relevant goal that users choose. Users receive a push notification twice a day to remind them of their strategy. When they record engagement in that strategy, they make progress on a visual ‘journey’ on the app and receive a song selected by a New Zealand celebrity or peer, accompanied by an encouraging message explaining how the song inspired them to feel, think and act positively. Engagement with the app is only expected to be ∼1 minute a day.

### Bro

Bro is a Chatbot app that delivers safety planning by engaging a young person in a dialogue. It was co-designed with rangatahi Māori to ensure that the app is accessible to Māori. Safety planning (41) is an evidence-based intervention (42–44) commonly used in face-to-face clinical settings for acute crisis presentations. Bro includes core features of safety planning including prompts to notice increasing distress, how to make the environment safe, and pre-prepared coping strategies, including social and crisis support as well as emotion regulation activities. A key feature is ‘the wall of strength’ that provides a reminder of aspects of a young person’s life that enhance wellbeing, provide meaning and purpose, and support people. Young people receive a push notification once a day to engage with Bro.

### LifeBuoy

LifeBuoy is an app co-designed in Australia with young people with lived experience of suicidal thoughts (45). LifeBuoy contains seven sequentially delivered modules derived from Dialectical Behaviour Therapy (DBT) and Acceptance and Commitment Therapy (ACT). Each module takes approximately ten minutes to complete, and teaches distress tolerance, emotion regulation, mindfulness skills, and values through interactive learning exercises to help young people develop strategies and skills for managing distress. There are no push notifications to engage participants back into the app, but within app self-monitoring trackers allow tracking of mood and time spent in self-care behaviours.

### My Mood Attention matched control app

My Mood is a daily mood-tracking tool. Users receive a push notification once a day to log their mood on a half-circle (with editable descriptors allowing young people to use their own words, based on a 0-10 scale) with an optional journal-style reflection feature. The app is intended for use in research settings as a control app, engaging young people in an app to control for the action of logging into an app and brief engagement.

### Outcomes

A summary of the measures and measurement schedule are in Table 1.

**Table 1.**
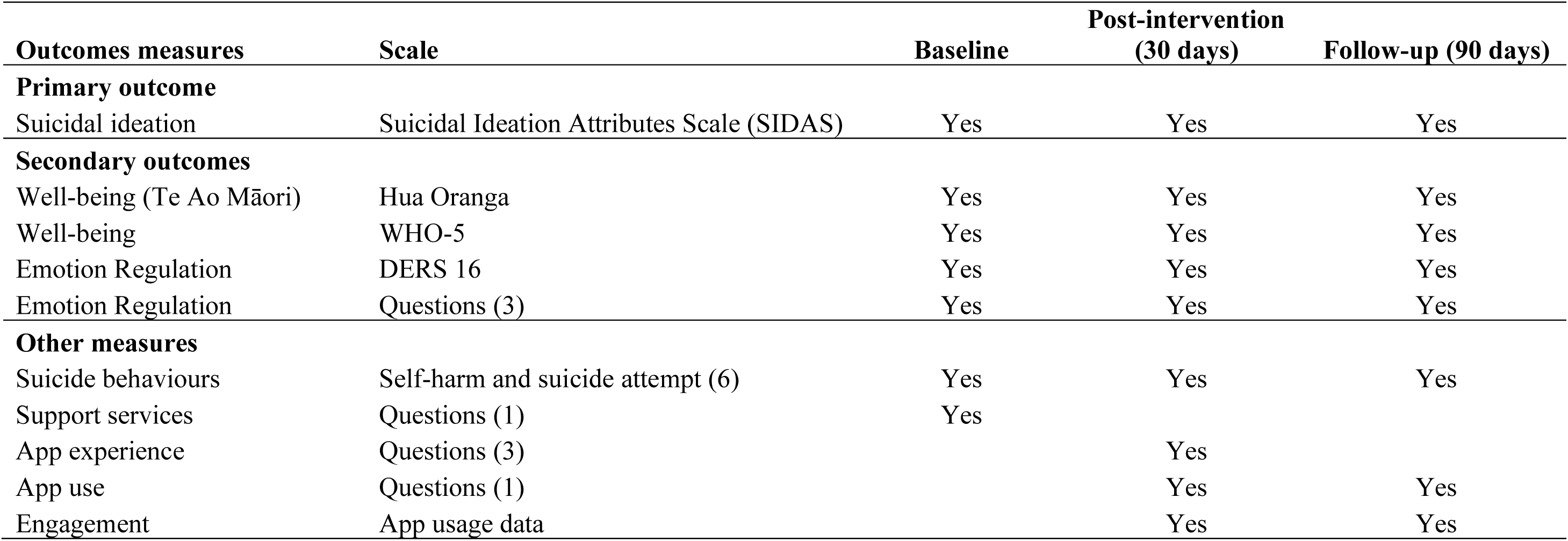
Summary of the primary and secondary outcomes, and other measures taken at the three assessment time points in the trial.

### Primary Outcome

#### Suicidal Ideation

The severity of suicidal thoughts will be measured by the Suicidal Ideation Attributes Scale (SIDAS (46)). The SIDAS maximum score is 50; scores of 21 or higher indicate more severe suicidal thoughts. The five questions pertain to the frequency of suicidal thoughts in the past month, controllability of suicidal thoughts, closeness to a suicide attempt, level of distress associated with the thoughts, and impact on daily functioning. Each item is assessed on an 11-point scale (0-10). Item two (controllability) is reverse-scored (46).

### Secondary Outcomes

The secondary outcome measures are:

### Mental Wellbeing

Mental wellbeing will be measured from a Te Ao Māori world view via the Hua Oranga (47) and World Health Organisation Wellbeing Index (WHO-5). The Hua Oranga comprises four items that measure wellbeing in terms of wairua (spiritual health), tinana (physical health), hinengaro (mental wellbeing) and whānau (family wellbeing), with each item rated from 1 (very bad) to 5 (extremely good); total scores range from 4 to 20. The WHO-5 has five items that assess wellbeing over the past two weeks on a six-point scale. The total raw score of 25 is multiplied by four to obtain a percentage score, with higher scores indicated greater wellbeing. It has is validated in young people (48) and has been used extensively in population-level studies in youth in Aotearoa New Zealand (4).

### Emotion regulation

Emotion regulation will be measured using the short-form16-item Difficulties in Emotion Regulation Scale (DERS-16), which maintains good construct validity, internal consistency and discriminative ability (49, 50) compared with the original DERS (51). The DERS-16 assesses acceptance of negative emotions, ability to engage in goal-directed behaviours when distressed, ability to control impulsive behaviours when distressed, access to emotion regulation strategies perceived as effective, and emotional clarity. Each item is rated on a 5-point Likert-type scale from 1 (almost never) to 5 (almost always). Total scores range from 16 to 80; higher scores reflect greater levels of emotion dysregulation.

A bespoke measure of emotion regulation skills for the purpose of developing a brief measure suitable for online trials was developed. It fills a gap in current measures by having behaviourally specific measure of capabilities via three items: “When I feel upset, I can calm myself down”; “When I am upset, I do healthy things to feel better e.g., reach out to someone who cares”; “Even when I feel upset, I know I’ll feel better later”; each is scored as 0 (not at all), 1 (sometimes) or 3 (all the time) with total scores ranging from 0 to 6.

### Acceptability

Acceptability of apps is rated via three bespoke questions: “Do you like the app?”; “This app is relevant to me and to people like me” both rated on a 5-point scale; and, “Do you plan to keep using it?” rated on a three point scale.

### Engagement

Engagement is measured via a single item question designed specifically for this trial: “I have used the app” with ratings of: Heaps (lots of days) (2); Sometimes (1); or, Not at all (0).

Engagement will also be measured via app usage, operationalised as time spent on the app, and the number of times key (app-relevant) events are logged (Lifebuoy: activity completion, mood monitor; Tune In: logged progress or no progress; changed journey; Bro: engaged in chat, viewed safety plan view, viewed wall of hope; My Mood: logged mood).

### Other measures collected at baseline

Participants are asked to provide their age, where they live (Australia or Aotearoa, New Zealand), their gender identity, ethnicity (s), prior use of counselling, therapy or medication for mental health problems health/well-being app use, motivation to engage in the trial, and lifetime history of suicide attempt or self-harm.

### Safety measures

To monitor the safety of participants, suicide attempt and self-harm are assessed by six questions developed specifically for this trial, specifically, if they have attempted suicide, and separately, engaged in self-harm in the past 30 days, for each provide the number of times, and indicate the severity of the most severe suicide attempt, and separately, the worst self-harm episode indicating that: ‘I didn’t need any medical care’, ‘I treated myself’ or ‘I went to the doctor’s or the hospital’.

### Qualitative interviews

After completion of the post-intervention (30-day) assessment, rangatahi Māori from each intervention app condition will be invited to participate in a 30-minute interview about the app that they have been using to explore acceptability and perceived impact. Interviews will be online and take place within four weeks of completing the post-intervention assessment to align with the primary endpoint and avoid memory degradation (based on the previous trial of Lifebuoy (35)). Interviews will be conducted by and for Māori within a kaupapa Māori framework (52, 53); they will open and close with karakia (Māori prayer/incantation) and an opportunity for whakawhanaungatanga (connection, relationship building). The Māori researcher will ensure that manaakitanga (support) is practised. The interviews will be audio recorded and transcribed verbatim.

### Sample Size

Sample size calculations are based on the primary hypothesis that will be tested for each site (Aotearoa New Zealand and Australia). This ensures adequate power to detect expected site-specific treatment effects. Based on an initial efficacy randomised trial of LifeBuoy (35), an effect size of 0.45 (Cohen’s d) is expected post-intervention (30 days) for suicidal ideation measured on the SIDAS for each app versus control. This would equate to differential changes of about 3.5/4 points on the SIDAS, which is considered a minimally clinically significant effect. Assuming this is the smallest effect size for any of the intervention apps relative to the control app, and assuming an attrition rate of 35% (as is typical in RCTs in mental health generally, including digital mental health trials (54)), a 0.50 correlation between scores on the repeated administrations of the SIDAS (a conservative estimate in case of a large correlation), and considering alpha adjustment for multiple comparisons (alpha = 0.01/3 intervention vs control comparisons = 0.003), a sample size for one site of n = 185 per trial arm (total site n = 740) will detect expected effect sizes with power = 0.90. A such, a total study sample size of 1,480 will be required.

The sample size for the interviews is up to 20 per app with the aim to have a balance in terms of the demographics of participants rather than representativeness. As such, adequacy of the sample size will be determined based on the richness and depth of the data to ensure there is sufficiency (55, 56).

### Randomisation

Participants will be allocated to study arms (1:1:1:1 ratio using a block design [four participants per block]), stratified by site (Aotearoa, New Zealand vs Australia) via a pre-generated randomisation list embedded in Tirohia. Using a blind-to-hypothesis approach, participants will not know whether the app they have been allocated is an intervention or the control app but will be informed they will be randomly assigned to one of four apps being evaluated for the self-management of suicidal ideation. This approach means that detection bias can be avoided, as all outcome measures are self-report.

### Blinding

All investigators and the trial statistician will remain blind to allocation. The trial manager and the data officer (administrative) who prepare data for the statistician and the Data Safety Monitoring Committee (DSMC) may become unblinded to participant allocation, and its potential influence is acknowledged.

The statistician will only receive app numbers (1 - 4) in the analysis dataset, without knowing which app each number represents. To prevent unblinding, the statistician will analyse primary outcome data before viewing and analysing app usage data, as usage patterns (time spent, logged events) differ by app and would reveal allocation.

### Data Collection and Management

Data will be collected using the Tirohia platform and stored securely on a University of Auckland Research Drive. Within Tirohia, study data will be collected in an identifiable form. Personally identifiable data will be stored separately from outcome data, which can be re-linked to data via the GUID. Re-identification is necessary to enact the safety protocol (i.e. if a participant needs to be contacted) as well as technical support and system administration.

All communications between the participant’s device and the Tirohia are encrypted. Tirohia resides on a secure University of Auckland server. Core system administration and trial manager access require two-factor authentication from any location, which is overseen by the University of Auckland Digital Services teams, who can detect and mitigate external threats in line with other University systems. The University has a well-developed cybersecurity policy, including monitoring, incident management, and, as appropriate, reporting.

The statistician will receive trial data using the data sharing process that aligns with the University of Auckland’s Research Data Management Policy (clause7). The deidentified data can be requested by other researchers for future research. A subset of our investigators will form a data request committee to review requests.

### Statistical Methods

Test of the primary hypothesis will involve directly comparing changes in suicidal ideation on the SIDAS from baseline to post-intervention (30-day), and from baseline to follow-up (90-day), for each of the individual intervention apps compared with that of the control app, using linear mixed models repeated measures analyses with maximum likelihood estimation, an appropriate covariance structure, and taking into account the effect of site. Given multiple comparisons, alpha will be adjusted (alpha = 0.01/3 intervention vs control comparisons = 0.003). Subsequent tests of the primary hypothesis will involve examining individual app intervention effects in New Zealand and Australia to understand the effects of apps developed for a different country, with similar analyses to those described being conducted for each site. A similar analytical approach will be used to test the secondary hypothesis related to change in mental well-being. To prevent missing data, we will follow up participants who do not initially complete outcome measures at post-intervention and follow-up. The linear mixed models approach is also capable of incorporating all available data, including participants with missing follow-up data points, under the missing-at-random assumption. Analyses will therefore accord with the intention-to-treat principle. Sensitivity analyses will be conducted where appropriate to determine whether trial outcomes are robust to different missing data assumptions (57). Where appropriate, to complement the intention-to-treat analyses, analyses will be repeated with an as-treated approach, including participants based on the app that they received. To examine emotional regulation as a potential mediator of suicidal ideation outcomes, linear mixed models with similar model specifications as previously described will be used to examine associations between each app and emotion regulation at each assessment, and between emotion regulation and suicidal ideation at each assessment, while accounting for baseline emotion regulation and suicidal ideation. Indirect effects will be estimated and evaluated using 95% bias-corrected and accelerated confidence intervals formed from 5000 bootstrapped samples. Engagement will be evaluated using the single question designed for the TIPS trial, and via descriptive statistics for app event log data. Acceptability will be evaluated: 1) quantitatively using responses to single questions about acceptability designed for the TIPS trial; and, 2) qualitatively with a main focus on rangatahi Māori via qualitative interviews (up to 60), which will be analysed using the reflexive Braun and Clarke approach (58–60) with patterns of meaning consistent with a Kaupapa Māori approach.

No interim analyses are planned. A Data Safety Monitoring Committee (DSMC) comprising those with statistical expertise, trials expertise, and youth mental health and digital health expertise will monitor recruitment, enrolment, retention (withdrawal and attrition), quality and completeness of data (examining extracts of research data that are logged from the online collection), and safety (including ability to halt the trial should there be concerns). They will also monitor the safety protocol (see below). The DSCM Charter is available on request.

### Safety protocol

Links to crisis helplines will be available via Tirohia, in the PIS and embedded within each outcome assessment.

If a participant’s SIDAS score exceeds 20 or self-harm is indicated at any assessment, an automated email is sent to both the participant (containing crisis helplines and an invitation for a call) and trial manager. The trial manager will follow up the same day to arrange an optional call with a study clinical psychologist, who is available on-call 9:00–17:00, seven days a week. This follow-up call is not a crisis response but ensures connection to natural supports and services available to them.

The trial protocol and statistical analysis plan can be obtained by emailing the corresponding author.

### Ethics and dissemination

Ethical approval was obtained from the Health and Disability Ethics Committees (HDEC; Ministry of Health, reference 2025 EXP 21500, 26 February 2025) and is registered with the Australian New Zealand Clinical Trials Registry (ACTRN12625000349448). Recruitment commenced on July 14, 2025, and is expected to be complete by June 2026. Results will be disseminated through peer-reviewed publications, the Te Ata Hāpara Suicide Prevention Research Centre’s and Black Dog Institute websites, media, to relevant policymakers and decision-makers, and at international scientific conferences. Protocol modifications will be reported to the trial sponsor, HDEC, ACTRN after convening the DSMC.

### Patient and Public Involvement

Rangatahi Māori and other young people with lived experience were central to the development of this research. The four apps were developed through co-design with young people. The Rangatahi Kairangahau Tīpu refined the study protocol. This provided critical input to the design of recruitment strategies and study materials to ensure they are engaging, safe, and culturally appropriate. The selection of outcome measures was informed by prior participatory research prioritising outcomes relevant to young people. This group will oversee the ongoing conduct of the trial and co-design a dissemination strategy to ensure that the findings reach youth and community audiences effectively.

## Discussion

This trial is a critical step in youth suicide prevention. To our knowledge, this is the first and one of the largest trials to test the efficacy of a range of apps designed to reduce suicidal ideation in young people. Crucially, it will generate important information on the generalisability and true scalability of these interventions in a global context. This information will inform the extent to which apps are culturally appropriate, with a particular focus on the Indigenous people of New Zealand, Māori.

There are some limitations. Firstly, there are concerns about fraudulent behaviour common to online trials (61). While SMS verification mitigates multiple entries and provides protection against automated bots, the trial relies on participants truthfully reporting eligibility criteria and age; there is no technical mechanism to ensure participants are aged between 16 and24. Mitigation strategies include automated and manual checks of new participant accounts and a one-time phone number verification process.

Secondly, recruitment methods vary across the two sites, Aotearoa New Zealand and Australia. Both sites will primarily use paid social media advertising. In Aotearoa New Zealand, recruitment will be augmented by printed posters displaying the QR code/URL placed in community youth venues, schools, and universities, using advertising co-designed by a Rōpu Kairangahau Tipu Māori (Māori youth advisory group). Co-designed content will also be shared through trusted partner organisations.

Finally, the SIDAS presents suicidal ideation as linear, when it is a highly fluctuating phenomenon. The online presentation logic means that failure to endorse the first item results in no data being collected for the primary outcome. Furthermore, while the DERS will capture coping skills, the outcomes were not co-designed, which has been shown to be critical for capturing relevant measures, e.g. for managing in-the-moment distress (62, 63).

## Author Statement

SH, TC, SF, MT, LM, HT, and QW conceptualised the research goals and aims and obtained the funding; NL, IM, LM, and SH managed project administration. SH, TC, SF, MT, LM, NL and QW developed the methodology. SH and NL drafted the manuscript, and all authors contributed to its review and editing, approved the version to be published and agree to be accountable for the work.

## Data Availability

All data produced in the present study are available upon reasonable request to the authors

## Acknowledgements

We are grateful for the contributions of the Rōpu Kairangahau Tipu Māori and the young people with lived experience who were involved in the co-design the app development and recruitment strategies. We also thank the governance group Te Pou Whanui and Te Ata Hāpara (Suicide Prevention Aotearoa), and Youthline, the Mental Health Foundation, and Voices of Hope.

